# Circulating transcriptional biomarkers serve as surrogates of disease progression in Charcot-Marie-Tooth 1A disease

**DOI:** 10.1101/2025.08.28.25334434

**Authors:** L. Linhoff, A. Leha, B. Ahmad, T. Linhoff, S. Wernick, K. Kummer, S. Fritzsch, E. Akova-Öztürk, B. Dräger, B. Schlotter-Weigel, S. Thiele, N. Garcia-Angarita, E. Greckl, L. Reinecke, M. Rossner, M.C. Walter, P. Young, M.W. Sereda

## Abstract

**Objective:** Charcot-Marie-Tooth disease type 1A (CMT1A), the most common inherited peripheral neuropathy, results from PMP22 gene duplication and presents with progressive motor and sensory deficits. Despite well-defined genetics, therapeutic development is hindered by the lack of sensitive clinical outcome assessments (CAO) given slow disease progression and variable severity. This study aimed to identify robust transcriptomic biomarkers from blood which may serve as surrogate outcome measures for monitoring disease progression.

**Methods:** A cohort of 139 genetically confirmed CMT1A patients was longitudinally assessed over 2 years using standardized clinical tests and blood RNA sequencing at 3 expert sites across Germany. Parallel analyses were conducted in a Pmp22-overexpressing rat model over 12 weeks. Differential expression analysis, cross-species comparisons, and integration with functional data were employed to identify candidate biomarkers associated with disease severity and progression.

**Results:** Over the two-year follow-up, we confirm deterioration of the Charcot-Marie-Tooth Neuropathy Score (CMTNSv2) (0.32 (95%-CI [0.05; 0.59]) per year (p = 0.019). Out of a pool of consistently upregulated disease markers in CMT1A patients and CMT rats (vs.controls) based on RNA Seq data sets at baseline, 9 regulated genes were validated via Real-Time Quantitative PCR (RT-PCR). Transcriptional expressions of Actb, E2F2, MFAP, SAMD14, SEMA5A and SPI1 show a significant increase over time, whereas UQCRB declined over time.

**Interpretation:** Transcriptomic profiling revealed several blood-derived mRNA candidates in CMT1A patients correlate with longitudinal changes in CAOs and overlap with findings from the CMT rat model. These markers may offer a minimally invasive, scalable tool for disease monitoring in future clinical trials.

## Introduction

Charcot-Marie-Tooth (CMT) disease is the most common inherited disorder of the peripheral nervous system, affecting 10–28 per 100,000 individuals^1^. CMT patients show slowly progressive motor and sensory neuropathy and present with distal limb weakness, sensory loss, and foot deformities. The onset of disease can be observed in childhood, but diagnosis is usually made in the second decade of life. No therapy is available^2^. The most prevalent demyelinating subtype CMT1A, caused by a duplication of the PMP22 gene on chromosome 17 (17p11.1-12), accounts for 90% of CMT1 cases and 40–50% of all CMT cases^3^. CMT1A does not reduce life expectancy but significant impairments in everyday life are depend on the severity of the disease which shows a high variability, even among monozygotic twins^4^. Histopathological features include dys- and demyelination, onion bulb formation, and secondary axonal loss^5^.

Multiple preclinical approaches to lower PMP22 expression in CMT1A have been performed including progesterone antagonists^6,7^, PXT3003^8^ and ASOs^9^. The most recent well powered large multicentered Phase III trial of the combinatory drug PXT3003 demonstrated a good safety profile^10^, however, the prespecified endpoints were not met. Despite these advances, CMT1A’s slow progression pose a massive challenge for clinical trials which require sensitive measures to detect subtle changes. CAOs, such as the CMTNSv2, Charcot-Marie-Tooth Pediatric Scale (CMTPeds), and the ONLS, often fail to capture treatment effects in short-term studies^11–14^. Failed trials, including those repurposing ascorbic acid, underscore the need for advanced diagnostic approaches^15,16^. In response to these limitations, Rasch-modified versions of the CMT Examination Score (CMTES-R) have demonstrated improved sensitivity to disease progression. A large longitudinal study of 1,209 patients with CMT1A found that CMTES-R significantly detected change over a two-year period (mean 0.59 points, p = 0.0004), with a 55% improvement in the standardized response mean (SRM) over the original CMTES^17^. Similarly, in CMT1B, caused by MPZ point mutations, a distinct subtype of demyelinating and axonal neuropathy, CMTES and CMTES-R scores also showed statistically significant progression over time, with the greatest sensitivity in patients with moderate disease severity^18^.

The integration of novel biomarkers is transforming the landscape of CMT research. Muscle MRI fat fraction has proven the most sensitive biomarker in assessing disease progression in different CMT subtypes to date^19–21^. Skin-derived mRNA transcripts may serve as severity and progression markers in CMT1A^22,23^. Plasma neurofilament light chain (NfL) levels correlate with disease severity in CMT subforms, and in a translational approach, NCAM1 and GDF15 were shown to be biomarkers of CMT in patients and mice ^22,24–26^, but longitudinal data is missing. “CMT rats,” which overexpress PMP22 approximately 1,6 fold, have demonstrated similar disease characteristics as CMT1A patients, including dysmyelination, onion bulb formation, axonal loss, muscle atrophy and a high clinical variability similar to patients^27,28^. Thus, rodent models may provide critical data for disease progression which may contribute to identify sensitive patient biomarkers.^22^

To ensure clinical applicability, the validation of transcriptomic markers in this study followed a structured, four-phase “fit-for-purpose” framework encompassing pre-validation, exploratory and advanced validation, and in-study performance assessment. We combined pre-validation and exploratory validation by integrating patient data with CMT rats. Following the exploratory-validation, and applying stringent statistical testing methods, we aimed to identify novel and robust surrogate markers and bridge preclinical and clinical findings.

## Methods

### Ethics and Approvals

The trial was approved by the ethics committee of the University Medical Center Göttingen, Germany (UMG) under application number 31/2/16, with an initial positive vote on July 28, 2016, and subsequent amendment approval on May 22, 2018. Approval by the Ethics Committee of the LMU Munich was obtained under application number 173-16, reference 45–14. All participants provided written informed consent, and experiments adhered to guidelines for the ethical use of human and animal subjects in research. Animal experiments were conducted in compliance with German animal welfare regulations (TierSchG §4 Abs. 3) and the European Communities Council Directive 2010/63/EU, with approvals from the MPI NAT (formerly Max Planck Institute for Experimental Medicine’s) institutional authorities and the Lower Saxony State Office for Consumer Protection and Food Safety (LAVES Niedersachsen, license number 16/2200).

### Patients CMT-NET cohort

Patients were recruited from the German CMT-NET network at three University based sites (Munich, Münster, Göttingen). A total of 139 genetically confirmed CMT1A patients (male and female) were included and evaluated three times over two years at CMT-NET study centers. Standardized clinical evaluations included the CMTNSv2, ONLS, six-minute walk test, ten-meter walk test, nine-hole peg test, and maximal voluntary isometric contraction (MVIC) testing (measuring fist grip, three-point grip, foot dorsiflexion, and plantar flexion), following established protocols^12,29^. All examiners were trained and SOPs implemented. Blood samples were collected at each visit and immediately stored in PaxGene tubes for RNA stabilization as described^22^.

### CMT Rat Cohort

The "CMT rat" (SD-Tg(Pmp22)Kan; RDG ID: 2312447) model, a heterozygote Sprague Dawley strain with overexpression of the Pmp22 gene^27^, was bred to a cohort of 150 animals. From these, 4–5 rats were selected by comparing their phenotype for analysis in subgroups of mildly and severely affected animals. Behavioral phenotyping, conducted in week four and every consecutive four weeks after, included grip strength assessments using a Newton meter with a T-bar attachment and inclined plate test^30^. At week six, blood was collected for analysis. At week eighteen, electrophysiological analysis was conducted and blood was collected followed by dissection for tissue collection. Femoral nerves were fixed and used for histological analysis of axonal counting, axon caliber size and myelin thickness on the light microscopic level^30^.

### Library Preparation and Data Analysis

RNA-seq libraries were generated using the QuantSeq 3′ mRNA-Seq Library Prep Kit (Lexogen) and sequenced on the Illumina NextSeq 500 platform. Raw reads were demultiplexed by index and subjected to quality control using FASTQC. Subsequent alignment to reference genomes (human: hg19; rat: rn6) was performed with the STAR aligner. Gene-level quantification relied on FeatureCounts, using Ensembl transcript annotations (human: release 75; rat: release 93). Differential expression analysis was conducted with the DESeq2 R package, applying stringent thresholds (adjusted p-value < 0.05; |log₂ fold change| ≥ 0.66) to identify transcripts of interest. Comparative analyses across patient and rat cohorts prioritized shared dysregulated genes, with particular attention to those implicated in disease severity. Functional enrichment and pathway mapping were conducted using Cytoscape and ClueGO to contextualize transcriptional alterations within biological networks.

### Cross-Species Comparative Analysis

To identify conserved molecular signatures between humans and rats, a cross-species homology mapping was conducted to enable direct comparison of differentially expressed genes (DEGs) between CMT1A patients and Pmp22-overexpressing rats. DEGs exhibiting concordant directional changes across species and statistical significance in the patient cohort were prioritized for downstream analyses. Longitudinal transcriptomic profiling of patient samples at the first and third clinical visit facilitated identification of temporally dynamic genes. For further refining genes of interest, intersecting DEGs across timepoints were used to define robust candidate targets. Subsequent filtering incorporated significance thresholds tailored to each dataset, with a conservative false discovery threshold of p < 0.05 applied to human data and a relaxed cutoff of p < 1.0 for the rat model, reflecting the exploratory nature of cross-species validation.

### Analysis across visits

RNA-seq data analysis of four patients across two timepoints was performed using Galaxy (accessed on: 1 August 2022; https://usegalaxy.eu/). The data was cleaned from adapters using Trim Galore. Quality control was performed utilizing FastQC and MultiQC. We analyzed the mean quality score, GC content per sequence, and quality per sequence. RNA-Star (Galaxy Version 2.6.0b-1) was used alongside the most recent genome reference consortium file (GRCh38). In the next step FeatureCounts (Galaxy Version 1.6.3) determined the quantity of genes expressed. For further downstream analysis DESeq2 (Galaxy Version 2.11.40.4) filtered significantly differentially expressed transcripts p-value < 0.05, |log_2_ fold change| ≥ 0.66).

Lastly over- and under-expressed genes were identified by comparing differently expressed genes in severe compared to mild in both visits. To more precisely find impactful genes only transcripts showing in both visits were used. To identify the biological processes involving these genes we used Cytoscape 3.8.11 and ClueGo with GO as pathway source. Afterwards networks containing significantly differentially expressed genes with |log_2_ fold change| ≥0.66 were created. For further analysis we selected genes with the highest fold change between v1 and v3.

### Clinical Parameters

CMT patients were classified as mild (CMTNS < 10), moderate (CMTNS 10-20) or severe (CMTNS > 20). Clinical parameters (CPs) at the first visit were compared between all severity levels using the Jonckheere-Terpstra test and between mild and severe cases as well as between cases and controls using Welch t-tests or Fisher’s exact test, as appropriate.

### Preparation of Gene Expression Data

Based on visual inspection, one raw measurement in Batch 1 appeared as an extreme outlier and was set to missing. Additionally, several samples from the second visit in Batch 2 showed consistently high values and were also excluded. ΔCt values were calculated for each probe as the difference between the Ct of the gene of interest and the mean Ct of the housekeeping genes (ΔCt = Ct_gene_ − mean(Ct_housekeeper_)). Gene-specific plate effects were evident upon inspection, suggesting systematic variation across plates. To correct this, plate normalization was performed by subtracting estimated plate effects. These effects were quantified using test patient data through linear mixed-effects models, with gene, plate, and their interaction as predictors. Gene-specific plate effects were derived as marginal means from these models and used to adjust the expression values accordingly.

### Genes/CPs Over Time

Slopes were calculated to summarize progression over time by fitting individual linear models for each gene or patient. For each gene linear mixed effect regression models were fit to the expression using time as a predictor and additionally controlling for age, sex, and BMI. Resulting model coefficients were reported with 95% confidence intervals. Additionally, models including CMT severity at baseline and its interaction with time were fit and marginal mean effects of time within the CMT severity classes were estimated and reported with 95% confidence intervals. P values were adjusted across genes and CMT severity classes using the Bonferroni correction.

### Group Comparisons of Gene Expression

Gene expression was compared between groups at the first visit. Gene-wise linear models were fit to the gene expression using group as predictor and additionally correcting for age, sex, and BMI. Resulting p-values were adjusted for multiple testing using the Bonferroni correction. For the comparison of the three ordinal groups mild-moderate-severe or controls-mild-severe p values from likelihood ratio tests are reported.

### RNA Extraction and RT-PCR Analysis

Pooling the interesting genes, eighteen candidates were chosen depending on cross-species expression, change over time and correlation with severity. For nine of the candidates total RNA was extracted from blood samples using the PaxGene Blood RNA Kit (Qiagen) according to the manufacturer’s protocol. RNA integrity and concentration were verified using Bioanalyzer (Agilent). Complementary DNA was synthesized using SuperScript III Reverse Transcriptase (Invitrogen) with oligo-dT and random primers. RT-PCR was performed using GoTaq qPCR Master Mix (Promega) on a Light Cycler 480 (Roche), with primer specificity confirmed by melting curve analysis. Statistical outliers were excluded using Grubbs’ test. RT-PCR analysis was performed comparing severity groups and a subset of controls. P values were adjusted using the Bonferroni correction.

### Modelling CPs at Baseline

Multi-variable regression models using the gene expression measurements together with age, sex, and body mass index (BMI) as predictors were fit to each clinical outcome. We used ordinal regression models for the ONLS Score Arm, logistic regression models for the extreme CMT severity (only mild and severe), and linear regression models otherwise. The resulting model coefficients are reported with 95% confidence intervals. P values were adjusted across the clinical outcomes using the Bonferroni correction. To also study effects in groups of (baseline) CMT severity (mild/moderate/severe), for each clinical outcome and each gene, separate models were fit where CMT severity and its interaction with gene expression were additionally included. Expected marginal means of the gene expression trend, within each CMT severity class, were estimated and reported with 95% confidence intervals. P values were adjusted across the clinical outcomes and genes using the Bonferroni correction.

### Modelling Slopes of CPs and gene expression

Analogous models were fit for CPs as outcome and gene expression as predictor using the slopes instead of the value at baseline. The significance level was set to alpha = 5% for all statistical tests. All analyses on the clinical data were performed with the statistical software R (version 4.2.3)

## Results

### Phenotyping of CMT Rats

In the Pmp22 transgenic rat model, disease variability was assessed by grip strength measurements, nerve conduction studies, and histological analysis^28^. Mildly and severely affected rats were stratified based on phenotyping every four weeks beginning with six weeks of age. After 18 weeks an electrophysiological exam was conducted, and the mice were sacrificed for histopathology (**Fig. 1 A**). Histological analysis of peripheral nerves was performed assessing myelination status and structural integrity across severity groups by Methylene blue and Gallas staining. G-ratio measurements, calculated as the ratio of the inner axonal perimeter to the total fiber perimeter, demonstrated hypermyelination for small caliber fibers and hypomyelination for large fibers comparing wildtype and CMT rats. Additionally, an increase in amyelinated fibers could be observed consistent with demyelination or impaired myelin maintenance as previously described (**Figure 1B** and data not shown)^31^. Grip strength was significantly different between WT and CMT rats including subgroups of mildly (MA) and severely (SA) affected animals (Grip strength at baseline ± SD; WT: 12.48 N ± 0.62 SD; MA: 9.61 N ± 0.77; SA: 4.16 N ± 2.7) (**Fig. 1C**) and differences were observed over the whole trial (**Fig. 1F**). Similarly, CMT rats exhibited significantly reduced CMAP amplitudes compared to WT, measured at the end of the trial, but no differences between severity groups (CMAP ± SD; WT: 3.63 mV ± 1.84; MA: 0.18 mV ± 0.22; SA: 0.16 mV ± 0.09) (**Fig. 1D**). As expected, nerve conduction velocity showed significant reduction in CMT rats (Nerve conduction velocity ± SD; WT: 56.65 m/s ± 7.34; MA: 18.4 m/s ± 4.99; SA: 10.4 m/s ± 3.9) (**Fig. 1E**). Testing on 25° and 35° inclined plates identified higher susceptibility in severely affected rats over time (**Fig. 1G-H**). All descriptive values are shown in **Table 1**.

**Figure 1:**
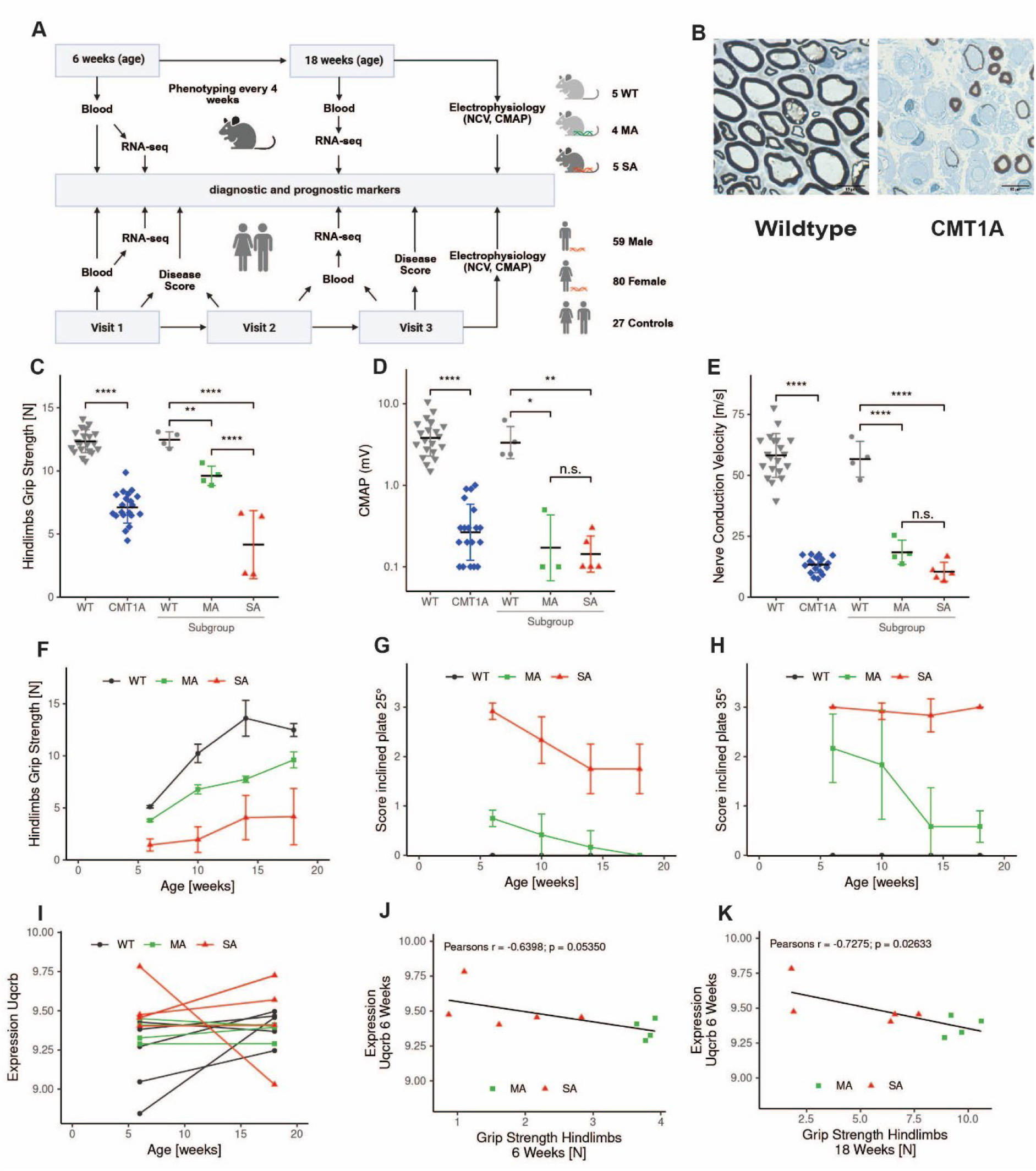
Study protocol in CMT rats and CMT1A patients. A: Flowchart of CMT rat phenotyping and assessment of clinical outcome measures in CMT1A patients and identification of transcriptional biomarkers from blood. *Pmp22^tg^* rats were phenotyped every four weeks starting at the age of six weeks until 18 weeks. At six weeks, based on the phenotype, 4 mildly and 5 severely affected animals were selected from a group of 150 animals. 5 WT age matched animals were used as control animals. Phenotype data is shown to show exemplary character of the selected animals. Blood was sampled at ages of 6 and 18W and electrophysiologic analysis was done at 18W. Blood samples at 6W were used for an RNAseq screen to identify disease markers. 80 female and 59 male CMT1A patients were recruited and examined over the course of two years in three visits. Each visit consisted of a detailed examination and blood sampling. 4 mildly and severely affected patients were selected to obtain an RNAseq data set using the blood sampled at visit 1. 27 healthy patients were recruited as controls. B: Peripheral nerve sections showing demyelination in CMT rats when compared to wildtype controls. C-E: Phenotyping using grip strength analysis, Inclined Plate 25° and 35° to identify severely and mildly affected CMT rats when compared to wildtypes; quantification with significant difference between severely, mildly affected rats and wildtype. F-H: Longitudinal phenotyping using Inclined 25°, 35° and grip strength. Significant separation of mildly, severely affected and wildtype animals I: Gene expression analysis over time of Uqcrb; significant increase from 6 to 18 weeks of age. J-K: Gene expression analysis and RT-PCR Validation of Uqcrb; Significant correlation of Hindlimb strength to Uqcrb at 6 and 18 weeks.

**Table 1.** Functional and Electrophysiological Characterization of Disease Severity in CMT rats.

| Parameter | Wildtype |  |  |  | Mildly Affected |  |  |  | Severely Affected |  |  |  |
| --- | --- | --- | --- | --- | --- | --- | --- | --- | --- | --- | --- | --- |
| Age (weeks) | 6 | 10 | 14 | 18 | 6 | 10 | 14 | 18 | 6 | 10 | 14 | 18 |
| Hindlimb Grip Strength (N) | 5.12<br>±<br>0.11 | 10.07<br>±<br>0.44 | 13.29<br>±<br>1.64 | 12.48<br>±<br>0.62 | 3.79<br>±<br>0.11 | 6.77<br>±<br>0.44 | 7.74<br>±<br>0.29 | 9.61<br>±<br>0.77 | 1.71<br>±<br>0.80 | 2.53<br>±<br>1.68 | 4.43<br>±<br>2.02 | 4.16<br>±<br>2.70 |
| CMAP Amplitude (mV) | 3.63 ± 1.84 |  |  |  | 0.18 ± 0.22 |  |  |  | 0.16 ± 0.09 |  |  |  |
| Nerve Conduction Velocity (m/s) | 56.65 ± 7.34 |  |  |  | 18.4 ± 4.99 |  |  |  | 10.4 ± 3.90 |  |  |  |
| Inclined Plate (25° & 35°) | Low susceptibility |  |  |  | Moderate susceptibility |  |  |  | High susceptibility |  |  |  |

### Genes of interest in CMT Rats

To identify candidates associated with disease severity, RNA-seq was conducted on whole blood samples obtained from CMT rats, comprising five mildly and four severely affected animals. This analysis yielded 248 DEGs between the two groups. To enhance translational relevance, an integrative comparison was performed with transcriptomic data from the human cohort, which revealed 45 overlapping gene candidates. Among these, Uqcrb exhibited the most robust association over time (**Fig. 1I**) and with disease severity in CMT rats after 6 weeks (**Fig. 1J**) and after 18 weeks (**Fig. 1K**). These cross-species candidates were subsequently prioritized for focused evaluation and validation within our human CMT1A cohort.

### Clinical Data of Patients

A total of 139 patients with CMT1A were recruited through the CMT-NET network. All patients were characterized at baseline and after two years. Whole blood samples from the two visits were utilized for RNA-Seq Analysis (**Fig. 1A**). The cohort exhibited a broad spectrum of disease severity, as indicated by the CMTNSv2, which ranged from mild (< 10), moderate (10-20) to severe (>20). The mean age at baseline (Visit 1) was 45 ± 11 years, ranging from 19 to 64 years. CMTNSv2 was estimated to change by 0.32 (95%-CI [0.05; 0.59]) per year (p = 0.019). The cohort comprised 80 (57.6%) female and 59 (42.4%) male patients. Follow-up data were available for 129 patients at Visit 2 (one year later) and 123 at Visit 3 (after two years). The ONLS total score only slightly rose from 3.5 ± 1.4 at baseline to 3.6 ± 1.6 at Visit 3 (Mean point change over 2 years: 0.068, p=0.388). The ONLS arm subscore also showed a slight increase from 1.6 ± 0.8 to 1.7 ± 0.9. However, the ONLS progression was statistically not significant.

Muscle strength assessments presented varying trends. Handgrip strength, measured by dynamometry, remained consistent at 23 ± 10 kg at Visits 1 and 2 before increasing to 25 ± 13 kg at Visit 3 (p = 0.051). Three-point pinch strength followed a similar pattern, remaining stable at 48 ± 26 N at baseline and showing only minor changes at subsequent visits (p = 0.089). Foot dorsiflexion strength exhibited a slight improvement, rising from 48 ± 46 N at Visit 1 to 53 ± 48 N at Visit 3 (p = 0.038). Meanwhile, plantar flexion strength fluctuated between visits, decreasing to 79 ± 58 N at Visit 2 before returning to 85 ± 63 N at Visit 3 (p = 0.067).

Gait assessments reflected a modest decline in mobility. The 10-Meter Walk Test (10MWT) recorded a slight increase in walking time, from 7.9 ± 2.7 s at baseline to 8.2 ± 3.1 s at Visit 3 (p = 0.029). Conversely, the 6-Minute Walk Test (6MWT) remained relatively stable, with distances of 435 ± 120 m at Visit 1, 444 ± 109 m at Visit 2, and 447 ± 120 m at Visit 3 (p = 0.074).

Electrophysiological assessments demonstrated subtle but important changes. CMAP declined from 4.7 ± 3.6 mV at baseline to 3.4 ± 2.2 mV at Visit 3 (p = 0.022). As expected, motor nerve conduction velocity (mNLG) remained unaltered, with values ranging from 22 ± 8 m/s at Visit 1 to 24 ± 7 m/s at Visit 2, and to 23 ± 6 m/s at Visit 3 (p = 0.058).

Quality-of-life assessments revealed little variation over time. The SF-36 physical component summary score remained stable, ranging from 37 ± 12 at Visit 1 to 38 ± 12 at Visit 2 and returning to 37 ± 12 at Visit 3 (p = 0.091). Similarly, the SF-36 mental component score showed no significant changes, remaining at 49 ± 10 at baseline, 48 ± 12 at Visit 2, and 49 ± 11 at Visit 3 (p = 0.083). Pain levels, as assessed by the Visual Analog Scale (VAS), demonstrated a slight reduction from 3.5 ± 2.6 at Visit 1 to 3.3 ± 2.6 at Visit 3 (p = 0.067).

Overall, patients with CMT1A experienced a gradual but measurable decline in functional and electrophysiological markers over the two-year period. While CMTNSv2 indicated worsening disease severity, other assessments, including gait function, muscle strength, and quality-of-life measures, remained relatively stable. Notably, electrophysiological markers revealed a decline in motor response amplitudes, although conduction velocities were preserved. These findings underscore the progressive nature of CMT1A in our cohort (**Table 2**).

**Table 2.** Cohort Demographics, Clinical outcome measures (COM) and parameters (CP)

| Parameter | V1 | V2 (1 year) | V3 (2 years) |
| --- | --- | --- | --- |
| Sample size (n) | 139 | 129 | 123 |
| Age (years) | 45 ± 11 | 46 ± 11 | 47 ± 11 |
| Sex, m/f (%) | 59 (42.4%) / 80 (57.6%) | 55 (42.6%) / 74 (57.4%) | 52 (42.3%) / 71 (57.7%) |
| Weight (kg) | 80 ± 19 | 80 ± 20 | 80 ± 20 |
| BMI | 28 ± 6 | 28 ± 6 | 28 ± 6 |
| 6MWT | 435 ± 120 | 444 ± 109 | 447 ± 120 |
| dML(ms) | 7.8 ± 2.2 | 7.4 ± 1.9 | 7.3 ± 1.9 |
| MSAP (mV) | 4.7 ± 3.6 | 2.8 ± 2.2 | 3.4 ± 2.2 |
| Grip Strength (N) | 23 ± 10 | 23 ± 11 | 25 ± 13 |
| 10MWT Zeit (s) | 7.9 ± 2.7 | 7.8 ± 2.7 | 8.2 ± 3.1 |
| CMTNSv2 | 16 ± 6 | 17 ± 6 | 17 ± 6 |
| ONLS | 3.5 ± 1.4 | 3.6 ± 1.5 | 3.6 ± 1.6 |

### Blood-Derived Transcriptomic Biomarkers Reflect Disease Progression in CMT1A patients and rats

To explore transcriptomic correlates of disease progression, we performed RNA-Seq analysis on whole blood samples collected longitudinally from a subset of eight male patients with genetically confirmed CMT1A, stratified into mildly and severely affected groups (n = 4 each, index patients) based on baseline CMTNS scores. Differential gene expression analysis revealed 876 transcripts with significant expression differences between severity groups. Cross-sectional comparison between initial and follow-up visits identified 30 transcripts consistently upregulated in severely affected individuals (**supplementary Fig. 1**). Integrative analysis with transcriptomic data from Pmp22 transgenic (Pmp22^tg) rats identified 45 overlapping genes, including canonical regulators such as Actb, SPI1, and UQCRB, thereby underscoring the translational relevance and bidirectional validity of the experimental design. From this intersection, a focused panel of nine genes was prioritized based on sustained differential expression over time and cross-species concordance and validated in the whole cohort via RTPCR excluding the index patients. Notably, Actb, E2F2, MFAP4, SAMD14, SEMA5A, and SPI1 demonstrated a significant longitudinal increase in expression, whereas UQCRB showed a significant decline (**Fig 2A-G**). Two additional candidates, RAB5B and TPK1, did not exhibit significant temporal variation (**Fig. 2 H,I**). The dynamics observed suggest that these genes may serve as accessible blood-based indicators of disease progression in CMT1A. To assess the diagnostic and prognostic utility of these candidates, we evaluated their associations with COAs while controlling for age, sex, and BMI. Linear mixed-effects modeling across all available visits revealed significant time effects for eight out of nine transcripts (Actb, E2F2, MFAP4, SAMD14, SEMA5A, SPI1, TPK1, and UQCRB). After Bonferroni correction for multiple comparisons, seven genes retained statistical significance (Actb, E2F2, MFAP4, SAMD14, SEMA5A, SPI1, and UQCRB), supporting their role as robust molecular indicators of disease progression.

**Figure 2:**
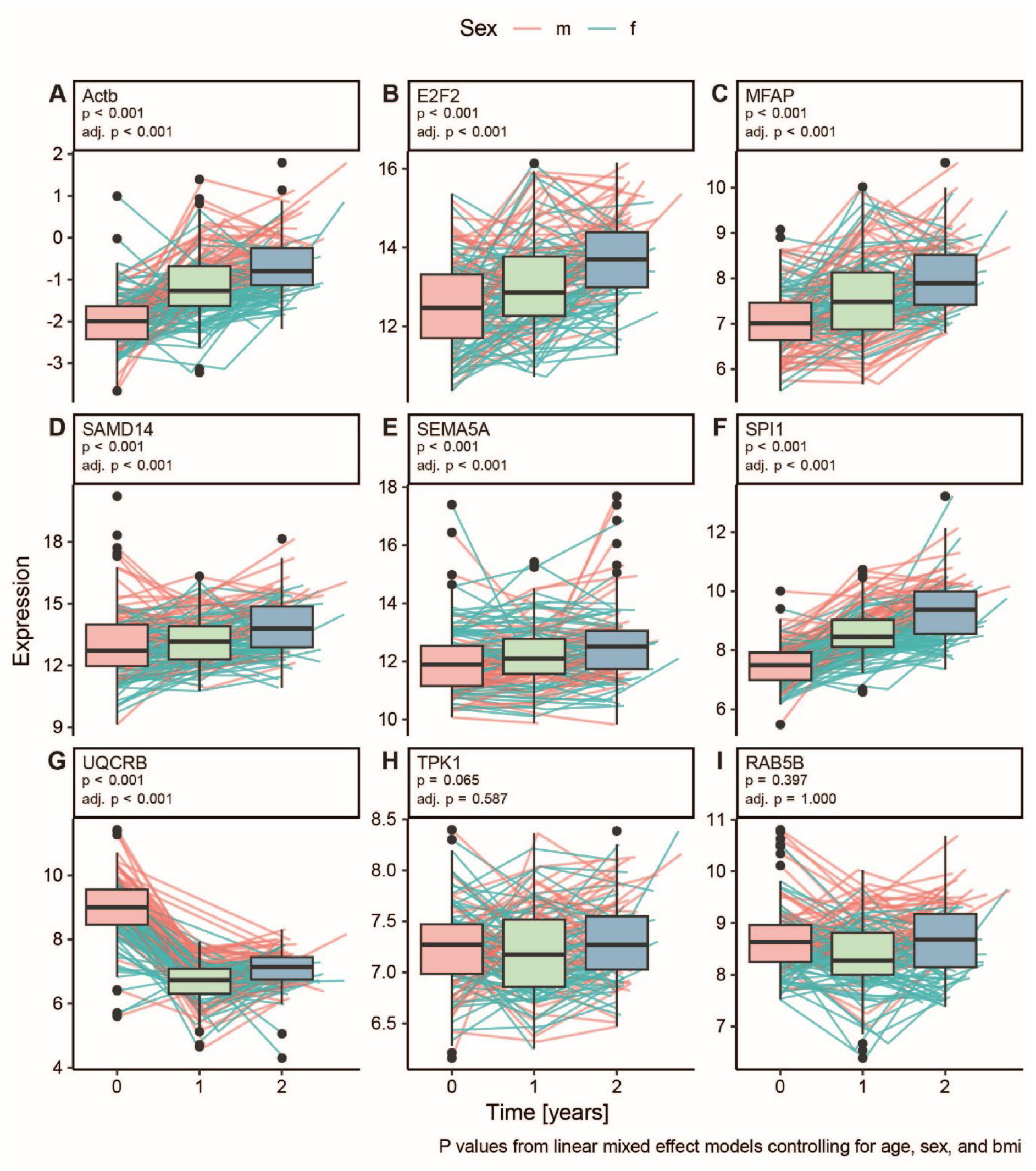
mRNA expression validated by RTPCR of 7 genes derived from blood in CMT1A patients progress over time. Nine potential biomarkers for CMT progression were assessed at three time points over 2 years in CMT1A patients. This Figure shows the expression changes across these three visits. Lines show expression changes for individual patients (red: male, blue: female). Significance was assessed in linear mixed effect regression models controlling for age, sex, and BMI; resulting raw and Bonferroni-adjusted p values are given. A-G: Progression of 7 significantly DEGs over time. H, I: Non-significantly altered expression over time

### Validation of transcriptional candidates in comparison with disease severity

In order to validate candidates from RNA Seq, we examined the most significant associations for gene expression by disease severity groups applying RT PCR. Significantly altered expressions for E2F2 (**Fig. 3A**), MFAP (**Fig. 3B**) and UQCRB (**Fig. 3C**) were observed when candidate expression in CMT1A patients were compared to controls, whereas we did not observe differences between severity groups as measured by the CMTNS (mild, moderate, severe). However, predictions using linear and ordinal regression modeling showed significant correlations (unadjusted) for all three candidates with CPs (CMAP and ONLS, Arm scores). After correction for multiple testing tendencies for E2F2 (**Fig. 3D**) and UQCRB (**Fig. 3F**) were observed. MFAP was not significant after correction (**Fig. 3E**).

**Figure 3:**
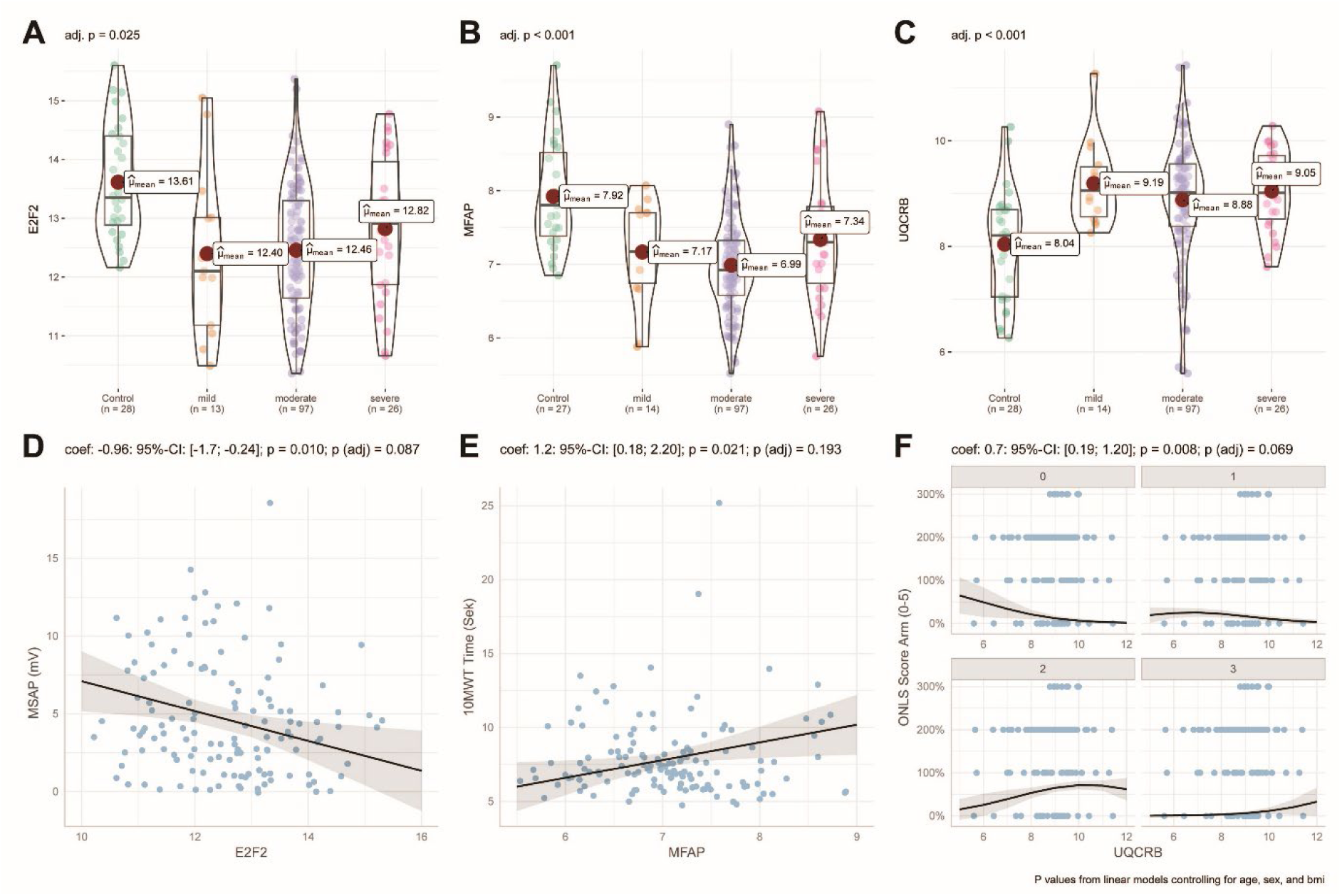
Correlation of transcriptional expression and COA and CPs at baseline. A-C: Significant differences of gene expression between CMT severity classes as measured by the CMTNS (mild, moderate, severe) and controls for the three biomarkers with the most significant associations for the gene expression using group as predictor. Although no differences were observed between severity groups, expression levels were significantly altered compared to controls. The data was corrected for age, sex, and BMI. D-F: Model predictions (lines) with 95% confidence intervals (shaded area) for the three most significant associations with CPs (CMAP and ONLS, Arm scores). Dots are individual measurements for patients. In (D, E) fits from linear models are shown, in (F) the fit from an ordinal regression model is shown, where the different sub-panels show the predicted probabilities for each level of the ONLS arms subscore (only levels 0-3 were present in the data). Linear and ordinal regression models showed significant unadjusted correlations for all three candidates; after correction for multiple testing, trends remained for E2F2 and UQCRB. P values were adjusted for multiple testing using the Bonferroni correction.

### Prognostic properties of candidates in patients

In the next step, we explored the prognostic properties of candidates. Regression models assessing the relationship between gene expressions at baseline and clinical progression slopes identified significant (unadjusted p < 0.05) associations to at least one of the 9 CPs in Actb (**Fig. 4A**), MFAP (**Fig. 4B-C**), RAB5B (**Fig. 4D-E**), SP1 (**Fig. 4F**), and UQCRB (**Fig. 4G**). After correction for multiple testing the associations between RAB5B and ΔONLS as well as SP1 and ΔSF36 (Physical) remained significant (**Fig. 4 D,F**).

**Figure 4.**
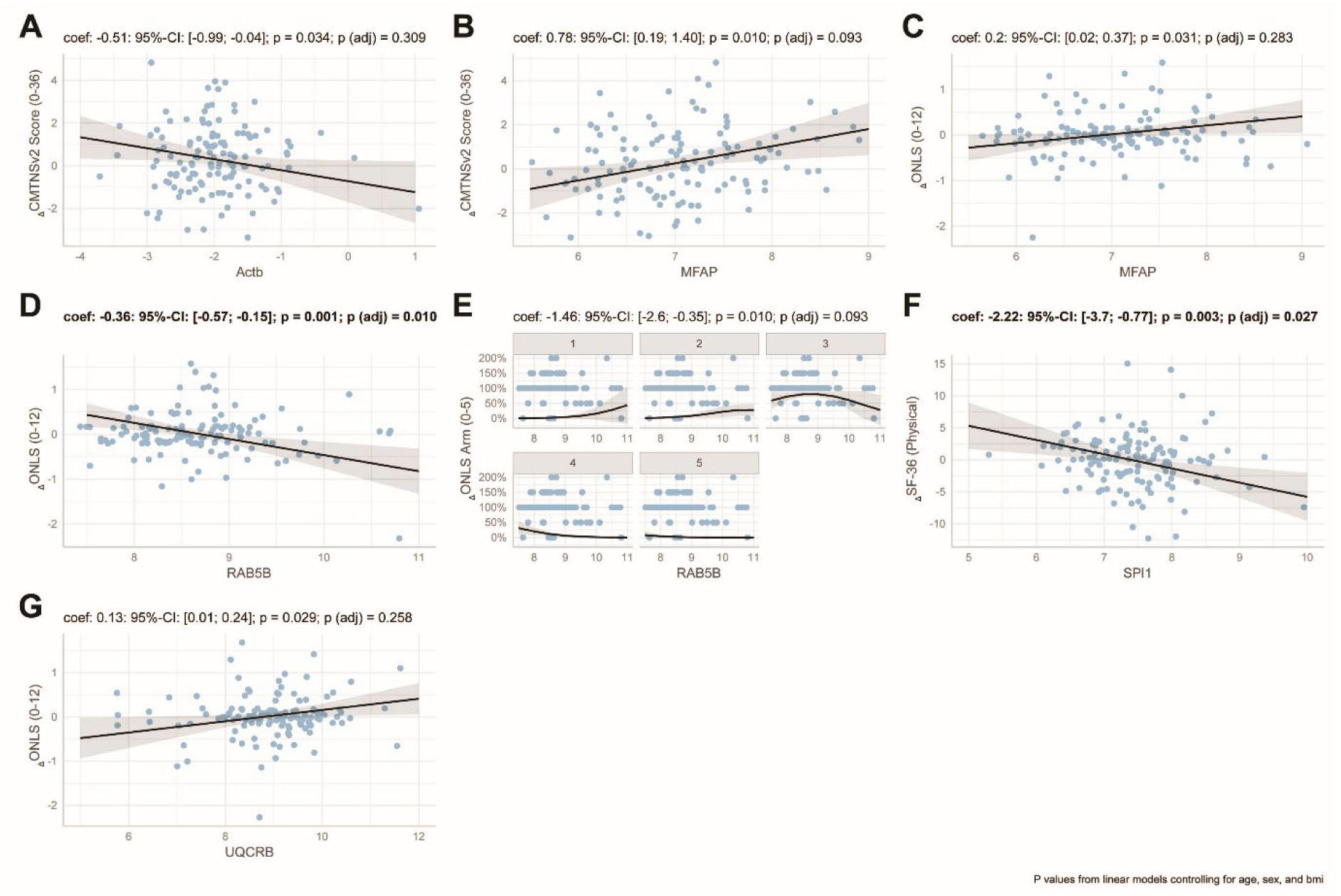
mRNA expression of candidate genes at baseline are prognostic for changes in CP and COA. A-G: Change of each clinical outcome assessment over two years was modeled using gene expression as predictor and controlling for age, sex, and BMI. This plot shows the model predictions (lines) with 95% confidence intervals (shaded area) for all associations with a (raw) p value < 0.05. Dots are individual measurements for patients. In panel (E) the fit from an ordinal regression model is shown, where the different sub-panels show the predicted probabilities for each level of the ONLS arms subscore (only levels 0-3 were present in the data). The other panels show fits from linear models. Regression analysis examining baseline gene expression in relation to clinical progression slopes revealed significant unadjusted associations (p < 0.05) for Actb, MFAP, RAB5B, SP1, and UQCRB with at least one of the nine CPs. After correction for multiple testing, significant associations remained between RAB5B and ΔONLS, as well as SP1 and ΔSF36 (Physical). P values were adjusted for multiple testing using the Bonferroni correction.

### Candidates as progression markers

Lastly, we assessed whether the change in candidate genes was associated with change in COAs. Overall correlations and subgroup analysis revealed Actb (**Fig. 5 A-B**), E2F2 (**Fig. 5C-D**), MFAP (**Fig. 5E-F**), RAB5B (**Fig. 5G**), SAMD14 (**Fig. 5H**), TPK1 (**Fig. 5I**), UQCRB (**Fig. 5J-K**) as correlating over time with mainly CMTNSv2, ONLS, CMAP, Dynamo foot lifting and SF-36 (unadjusted). After correcting for multiple testing, the associations of RAB5B with ONLS and MFAP with ONLS remained significant (**Fig. 5 F,G**) .

**Figure 5:**
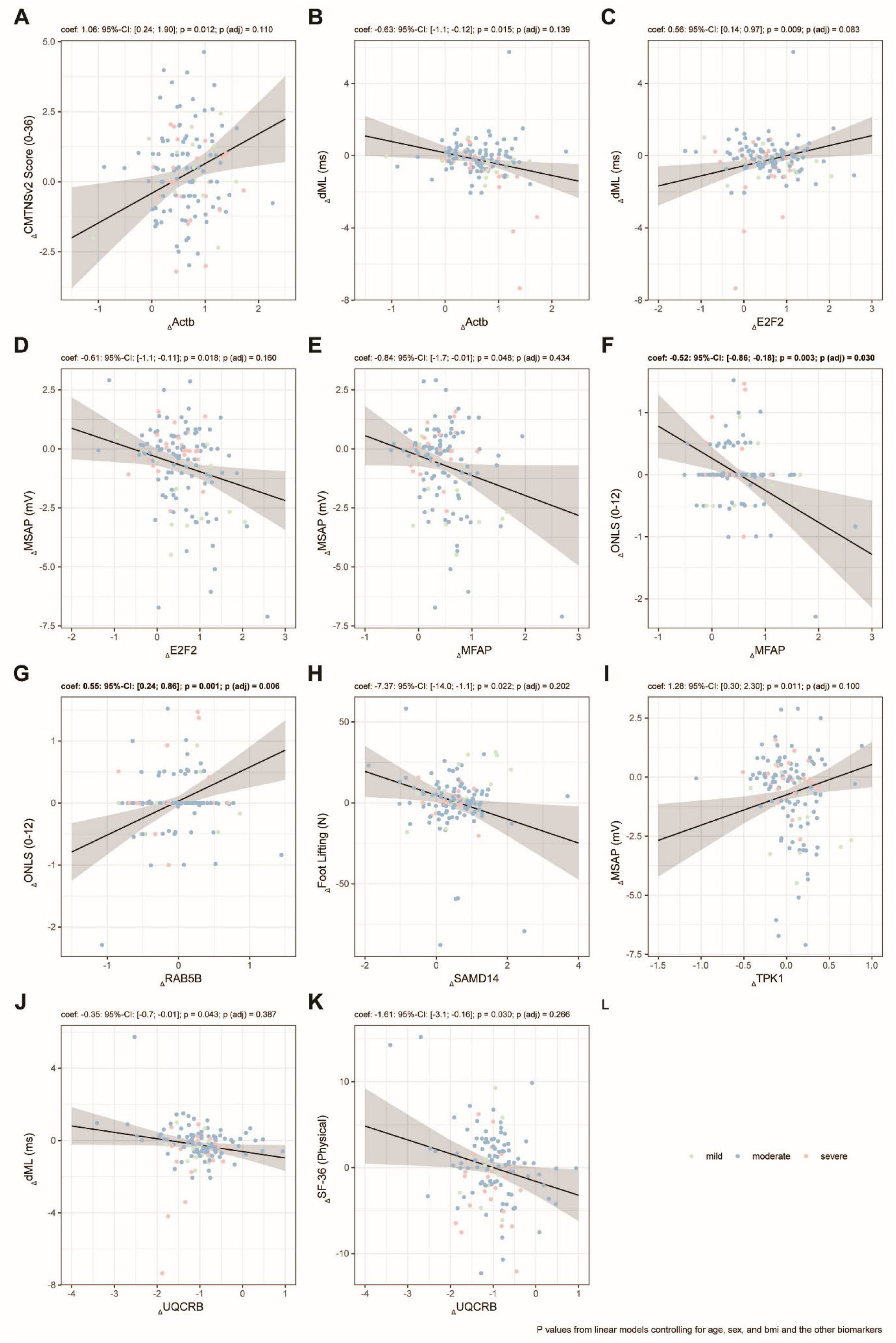
Correlations between changes in potential biomarkers and changes in CPs and outcome assessments. A-K: Change of each clinical outcome measure over two years was modeled using change in gene expression as predictor and controlling for age, sex, and BMI. This plot shows the model predictions (lines) with 95% confidence intervals (shaded area) for all associations with a raw p value < 0.05. Dots are individual measurements for patients. Overall correlations and subgroup analyses identified Actb, E2F2, MFAP, RAB5B, SAMD14, TPK1, and UQCRB as showing longitudinal associations— primarily with CMTNSv2, ONLS, electrophysiology, Dynamo foot lifting, and SF-36 scores (unadjusted). After correction for multiple testing, significant associations persisted between RAB5B and ONLS, and between MFAP and ONLS. P values were adjusted for multiple testing using the Bonferroni correction.

Divided into severity groups significant unadjusted correlations can be seen in mildly affected CMT1A patients for SAMD14 (**Fig. 6H**), moderately affected for E2F2 (**Fig. 6D**), MFAP (**Fig. 6E-F**), SAMD14 (**Fig. 6H**) and UQCRB (**Fig. 6J-K**) and for severely affected Actb (**Fig. 6B**) and UQCRB (**Fig. 6J**). Mildly and moderately affected patients mainly correlate with COAs, severely affected patients correlate with electrophysiological measures (CMAP; unadjusted), possibly reflecting axonal loss. Correcting for multiple testing, only the association between ONLS with MFAP in the moderate group (**Fig. 6F**) and dML with Actb stayed significant in the severely affected group (**Fig. 6B**).

**Figure 6.**
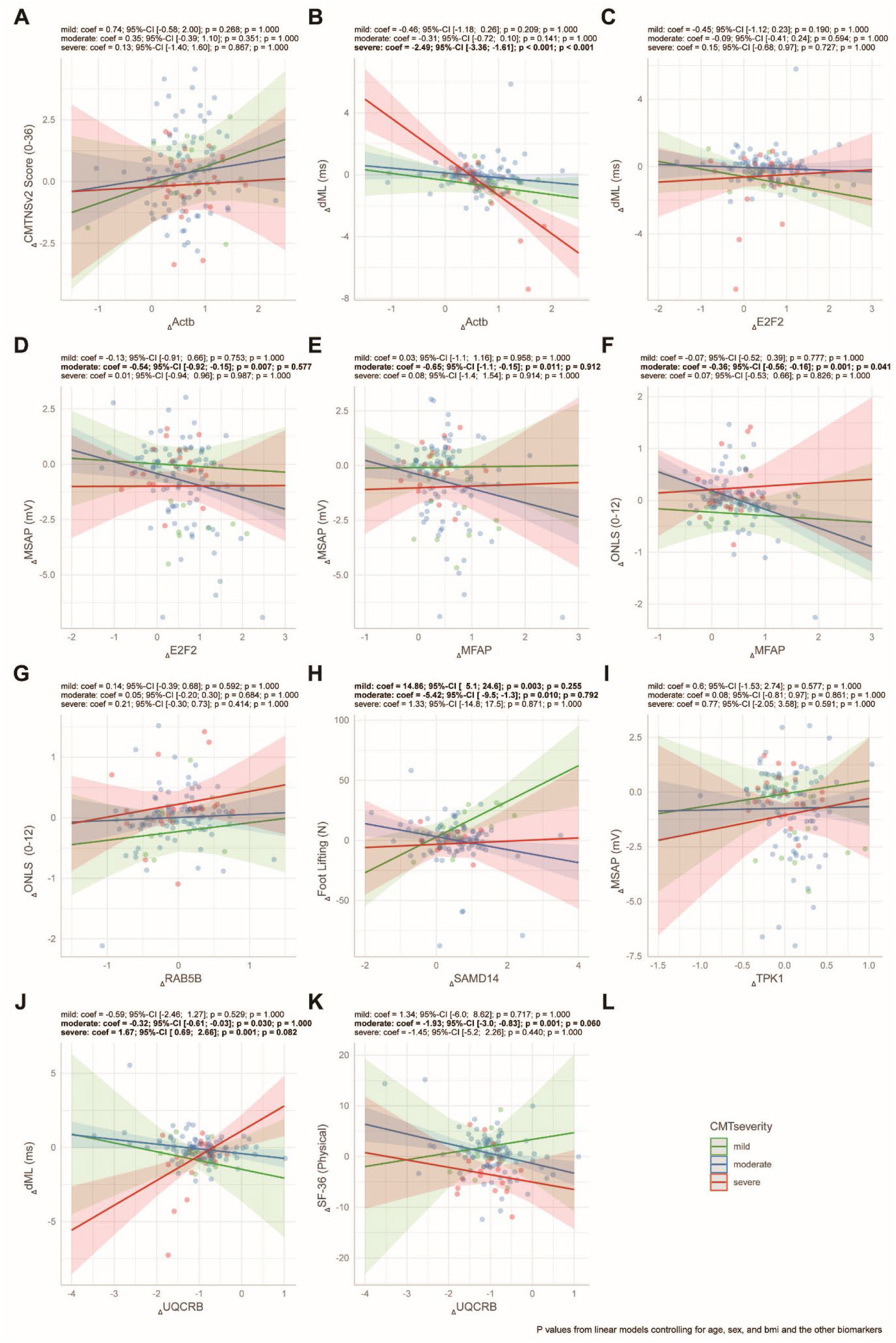
Correlation between changes in transcriptional expression and changes in CPs. A-K: For each potential biomarker the change of each clinical outcome measure over two years was modeled using the change in gene expression in interaction with baseline CMT severity as predictor and controlling for age, sex, and BMI. This plot shows the marginal means of the effects of the change in gene expression in each severity class (lines) with 95% confidence intervals (shaded area) for all associations with a (raw) p value < 0.05 in at least one of the severity classes. Dots are individual measurements for patients. When stratified by severity, significant unadjusted correlations were observed for SAMD14 in mildly affected individuals; E2F2, MFAP, SAMD14, and UQCRB in the moderately affected group; and Actb and UQCRB in severely affected patients. After correction for multiple testing, only the association between ONLS and MFAP in the moderate group remained significant. P values were adjusted for multiple testing using the Bonferroni correction.

## Discussion

In this study, we aimed to investigate whether blood-based transcript levels of selected candidate genes may serve as surrogate biomarkers for prognosis, disease severity, and progression in CMT1A. To address this, we conducted two complementary and parallel trials: one in a large cohort of genetically confirmed CMT1A patients, and the other in an established preclinical model, the Pmp22 transgenic (CMT) rat. This translational approach follows a similar framework to prior multinational studies that successfully identified skin biopsy–based transcriptomic signatures^22^. In our clinical trial, 139 CMT1A patients and 27 healthy controls were recruited across CMT-NET centers and evaluated at three time points over a two-year period. In parallel, a cohort of 150 adult CMT rats underwent behavioral phenotyping and molecular profiling over twelve weeks. Both human and animal cohorts demonstrated the expected features of slow disease progression and marked clinical heterogeneity. Alongside standard clinical outcome assessments (COAs) such as the CMTNSv2, we applied an extended battery of clinical and electrophysiological parameters, including the 10MWT, the 9HPT, and selected nerve conduction parameters. Following the fit-for-purpose framework, our study combines pre-validation and exploratory validation. As previously shown in CMT rat and patient derived skin biopsies, the overlap between human and rodent models is a strong tool in the search for biomarkers ^22^. To identify the most relevant transcripts, we first examined changes over time. **Combined with the overlap between rats and humans, we found nine genes of interest, of which seven (e.g. Actb, E2F2, MFAP, SAMD14, SEMA5A, SPI1 and UQCRB) showed significant alterations over time.**

To further demonstrate clinical relevance of our transcriptional alterations of the seven candidates, we performed statistical analysis using mixed models, with rigid adjustment for multiple testing. We first asked the question if the candidates are altered regarding severity groups. Comparing the gene expression between control and different CMT1A groups, we saw several significantly different genes between controls and CMT1A patients, but no differences when stratified by disease severity. However, individual transcripts such as E2F2 and MFAP4 demonstrated significant correlations with CPs or COAs at baseline, including the 10MWT and CMTNSv2. After adjustment for multiple testing there were only tendencies observed for E2F2.

We next assessed whether baseline expression levels of selected transcripts may predict clinical progression over time. **While five of the candidates (e.g. Actb, MFAP, RAB5B, SPI1 and UQCRB) were associated with clinical progression, only two candidates, MFAP4 and RAB5B, showed significant prognostic value, correlating with longitudinal changes in CMTNSv2.** When adjusted for multiple testing associations between MFAP with ONLS and RAB5B with ONLS were present. This result should be interpreted cautiously, as we saw no significant increase in ONLS in the timespan. Additionally, expression of SPI1 (PU.1) was found to correlate with patient-reported quality of life (SF-36). These findings suggest that the prognostic relevance of gene expression may vary across disease stages, with certain candidates, particularly MFAP and RAB5B, showing potential as severity-dependent biomarkers in CMT. Establishing robust biomarkers encompasses different stages of validation.

Why are multimodal assessments necessary? Given the slow progression of CMT1A, traditional COAs often fail to detect meaningful changes over short periods, limiting their utility in clinical trials. Established tools, such as the CMTNSv2 and the ONLS, have been refined but still lack the sensitivity needed to capture subtle disease progression^32^. Combining blood biomarkers with advanced imaging modalities and functional assessments may offer a comprehensive approach to addressing this gap. Muscle MRI, particularly fat-fraction analysis, has emerged as a robust tool for monitoring disease progression. Studies have demonstrated its sensitivity in detecting intramuscular fat accumulation over time, correlating with functional decline^19^. Integrating MRI findings with blood-based biomarkers, such as mRNA transcripts or NfL levels, could provide a multidimensional perspective on disease progression and treatment response. Functional outcome measures, such as the CMT Functional Outcome Measure (CMT-FOM) and the CMTPedS, further complement these approaches by capturing changes in mobility, dexterity, and quality of life. Recent validation efforts have established these tools as reliable metrics for assessing disease burden and therapeutic efficacy, also at early disease stages^14,33,34^. We note that only early intervention with NRG1 in Pmp22tg rats normalized PI3K-AKT/MEK-ERK signaling and prevented axonal loss, making early markers of disease progression essential^31^. Although our study here focused on adult patients, the use of Pmp22tg rats has been helpful in biomarker development for CMT1A at different disease stages (baseline, progression, prognostic). We note that CMT rats recapitulate key pathological features observed in patients, including dysmyelination, axonal loss, and motor deficits^25,27^. Behavioral phenotyping and transcriptomic analyses in these models have provided insights into disease mechanisms, identified potential molecular targets and previously contributed to biomarker identification in 2 previous studies^22,28^.

From the 7 candidate biomarkers identified in this study, we observed consistent alterations in transcripts that in some cases align with known molecular pathways implicated in peripheral nerve pathology. Actb encodes β-actin, a ubiquitous cytoskeletal protein and a widely used housekeeping gene due to its consistent high expression across cell types, including peripheral blood. E2F2 (E2F transcription factor 2) regulates cell cycle progression and proliferation. To date, no studies have reported altered E2F2 expression in the blood of CMT1A patients, nor has it been identified as a differentially expressed gene in CMT1A-related transcriptomic datasets. MFAP4, encoding microfibril-associated glycoprotein 4, is involved in extracellular matrix organization and elastin fiber maintenance. It is located at chromosome 17p11.2—within the Smith–Magenis syndrome deletion region, which is reciprocal to the 17p12 duplication observed in CMT1A. The typical PMP22 duplication in CMT1A does not consistently encompass MFAP4, although in cases where it does, it could serve as a modifier. While MFAP4 is expressed by mesenchymal cells and macrophages (notably serving as a macrophage-specific promoter in zebrafish)^35^, no human studies have examined MFAP4 mRNA levels in CMT1A blood^36^. SAMD14 (Sterile Alpha Motif Domain 14) is primarily expressed in hematopoietic stem and erythroid progenitor cells and is upregulated during stress erythropoiesis. Its baseline expression in peripheral blood is low, and there is no evidence of altered SAMD14 expression in CMT1A. Given that hematological stress is not a typical feature of CMT1A, its expression levels in patient blood are not linked to CMT1A. SEMA5A (Semaphorin 5A) plays roles in axon guidance and immune modulation and is expressed by Schwann cells during peripheral nerve myelination. It is regulated by Egr2/Krox20 and downregulated following nerve injury, suggesting a potential role in maintaining axon–Schwann cell integrity^37^. Although no transcriptomic studies have evaluated SEMA5A levels in CMT1A blood, its involvement in peripheral nerve biology makes it a gene of interest. Genetic studies have also reported rare duplications involving SEMA5A in patients with inherited neuropathies, including one CMT case, indicating that gene dosage variations could contribute to disease phenotypes, albeit rarely^38^. PU.1 (SPI1) is a critical transcription factor in myeloid lineage differentiation, influencing the development and function of monocytes, macrophages, and granulocytes^39^. Its dosage regulates immune cell fate and function, and it controls numerous macrophage-specific genes involved in antigen presentation and cytokine production. Dysregulated PU.1 expression could potentially exacerbate inflammation in CMT1A, where low-grade chronic immune activation in peripheral nerves may contribute to disease progression. RAB5B, a Rab GTPase, belongs to a superfamily of Rab, which play a role in endosomal trafficking. While demyelinating conditions remain unstudied, it may indirectly influence CMT1A pathology. No expression data is currently available for RAB5B in CMT1A patient blood. UQCRB encodes a mitochondrial complex III subunit and is expressed in all nucleated cells, including those in peripheral blood. There are no reports assessing UQCRB mRNA levels in the blood of individuals with CMT1A.

Why can whole-blood transcriptomic profiles reflect neuropathy? Although unknown in CMT patients, PNS injury and degeneration engage innate and adaptive immunity, releasing damage-associated molecular patterns (DAMPs) that activate immune receptors on circulating cells^40^. This provokes an inflammatory cascade: immune cells (e.g., monocytes, macrophages, T cells) are recruited and transcriptionally activated to clear debris and orchestrate repair^41,42^. Indeed, gene expression profiling of blood leukocytes in neuropathy patients reveals upregulation of inflammation- and stress-related genes (e.g., FOS, PTGS2, MMP9) that track with disease presence and severity^43^. Immune-mediated mechanisms are central in many neuropathies, creating measurable blood biomarkers of disease activity. Pro-inflammatory cytokines such as tumor necrosis factor-α (TNF-α) and interleukin-6 (IL-6) are elevated in patients with peripheral neuropathy and associate with more severe nerve dysfunction in diabetes mellitus^40,44^. For example, higher TNF-α levels correlate with slowed nerve conduction (reduced nerve velocity)^44^, and circulating IL-6 and IL-1 receptor antagonist concentrations rise in proportion to neuropathy severity scores^40^. Consistent with this, blood transcriptomic studies of Guillain–Barré syndrome (an autoimmune neuropathy) demonstrate heightened expression of immune-pathway genes, with MMP9 upregulation being a notable example. Additionally, MMP9 levels correlate closely with clinical severity in GBS and its animal models, underscoring how peripheral nerve damage drives systemic gene expression changes^43^. It remains unclear, why blood biomarkers may reflect localized nerve pathology, especially in early or mild disease stages of hereditary neuropathy like CMT. Therefore, variability in expression due to genetic, environmental, and physiological factors necessitates rigorous standardization in sample collection, storage, and analysis. Despite these challenges, the identification of a candidate gene panel for blood-based transcriptomic monitoring may represent an important advancement toward objective, scalable tools for CMT1A assessment. Our results warrant further validation in independent cohorts and clinical trials, e.g. the integration of transcriptomics into multimodal biomarker strategies alongside imaging and functional metrics.

Why do diagnostic, progression, or predictive markers not serve as severity markers? Neuropathies involve multiple mechanisms (e.g. metabolic imbalance, ischemia, inflammation, demyelination, axonal loss). A single biomarker usually captures only one aspect of this complex pathology. Additionally, nerve damage often has threshold effects – for example, a small loss of fibers might cause noticeable neuropathic pain, while further loss leads to numbness and may paradoxically decrease as nerves are completely lost. Such non-linear relationships mean a biomarker might increase early in disease but plateau later, breaking a simple severity correlation. Biomarker levels can vary widely between individuals due to genetic, environmental, or co-morbid factors. Neurofilament levels, for instance, vary significantly between individuals e.g. higher with age, even though they remain stable within the same individual^45^. This variability challenges defining a one-size-fits-all level that corresponds to mild vs. severe neuropathy across different patients. A moderately elevated cytokine or protein might signify severe disease in one person but only mild changes in another, depending on each person’s baseline. Biomarkers often reflect active disease processes more than cumulative damage. A patient with long-standing neuropathy may have severe permanent deficits, but if their disease is in a burnout or chronic stable phase, inflammatory or injury markers can return to normal. For instance, NfL is usually elevated when nerves are actively being injured (ongoing axonal degeneration); if a patient’s neuropathy has stabilized NfL may normalize even though the patient remains severely weak from past damage. In CIDP^46^, studies found no clear correlation between NfL level and current disability severity in treated or long-term patients, even though high NfL did predict future deterioration in some cases^47^. In short, biomarkers often capture the rate of damage, not the total damage accrued.

The integration of multimodal biomarkers and COMs represents a paradigm shift in CMT1A research. Future studies may comprise longitudinal studies combining blood transcriptomics, imaging, and functional assessments will be critical for validating these tools and establishing their clinical utility. The Pmp22tg rat model remains a key asset for bridging preclinical findings with human data, especially when focusing on overlapping gene expression patterns and functional correlations. To ensure the successful translation of these findings, future efforts should focus on standardizing protocols across centers and addressing the variability inherent in biomarker expression. Collaborative initiatives, such as the ACT-CMT study, provide a framework for achieving these goals by promoting rigorous validation and international collaboration^33^. Addressing the limitations of both animal models and human biomarker studies will be essential to improve the predictive value and applicability of these tools. In conclusion, this study identified several blood-derived transcriptomic candidates with potential diagnostic, prognostic, and severity-marker utility in CMT1A. These findings, supported by overlapping with animal model data, highlight the translational relevance of blood-based biomarkers for monitoring disease progression. The strengths of this approach include accessibility, minimal invasiveness, and compatibility with longitudinal study designs. However, limitations such as indirect correlation with nerve pathology and variability in expression underscore the need for standardized protocols and multimodal integration. Together, these results, derived from the largest and longest longitudinal CMT1A trial conducted in Germany, support the continued development of blood biomarkers as part of a comprehensive framework to enhance clinical trial sensitivity and therapeutic monitoring in CMT1A.

## Data Availability

All data produced in the present study are available upon reasonable request to the authors

https://www.clinicaltrials.gov/study/NCT06203093

## Acknowledgments

We extend our sincere gratitude to Beate Veith and Christian Wieczorek for their expert execution of animal phenotyping and histology, and to the CMT-NET Study Centers for their invaluable contributions to patient recruitment and clinical assessments. We also acknowledge Nirmal Kannaiyan at LMU Munich for their proficient handling of RNA sequencing and data processing. This study was made possible through the dedicated participation of 139 patients and the comprehensive analysis of the corresponding animal models over a two-year period (CMT-NET, CMT-MOD).

## Author Contributions

Conception and Design of the work (M.W.S., M.R., M.C.W., P.Y.), Acquisition of the data (K.K., M.C.W., P.Y., M.W.S.), Analysis and interpretation of the data (A.L., L.L., B.A., M.C.W., M.W.S.), Drafting the manuscript (M.W.S.), Revising it critically for important intellectual content (All Authors), Final approval of the version to be published (All Authors), Accountable for all aspects of the work (M.W.S). B.A., L.L. and A.L. have full access to all the data in the study and take responsibility for the integrity of the data and the accuracy of the data analysis.

