## Supplemental Figure 1 for "Circulating transcriptional biomarkers serve as surrogates of disease progression in Charcot-Marie-Tooth 1A disease"

**Supplementary Figures:**

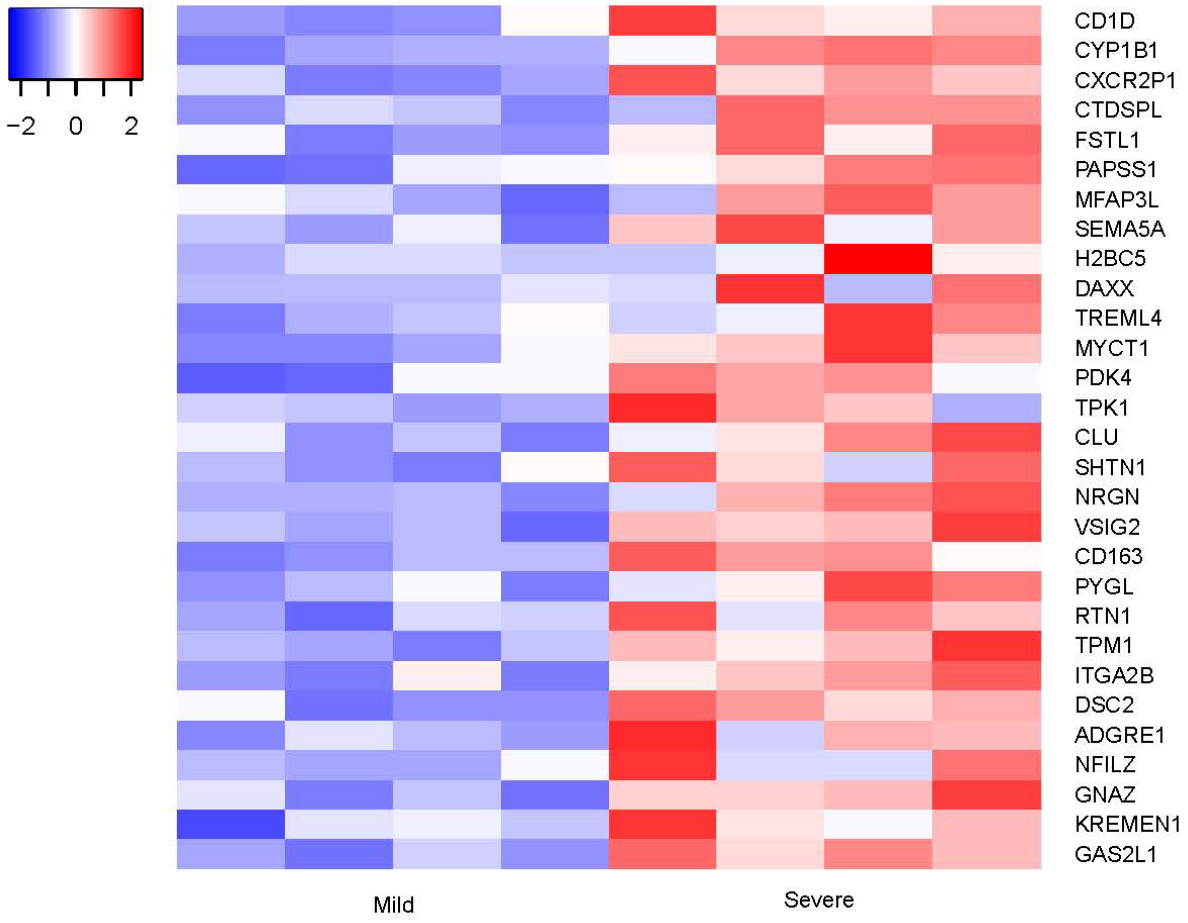


**Supplementary Figure 1 Differential gene expression in patients with mild and severe phenotypes.** Heatmap representation of selected differentially expressed genes between CMT patients stratified by clinical severity (Mild vs. Severe). Each row corresponds to an individual gene, and each column to a patient sample. Expression values were normalized and scaled by row to z-scores, with blue indicating relative downregulation and red indicating relative upregulation. The color scale ranges from –2 (low expression) to +2 (high expression). Genes shown were identified as significantly dysregulated between mild and severe CMT consistent over the two visits, highlighting molecular signatures associated with disease severity.
